# Identification of Metabolic Pathways Altered in Thyroid Cancer Progression and Metastasis

**DOI:** 10.1101/2023.09.24.23296027

**Authors:** Luís Jesuino de Oliveira Andrade, Luisa Correia Matos de Oliveira, Gabriela Correia Matos de Oliveira, Alcina Maria Vinhaes Bittencourt, Catharina Peixoto Silva, Luís Matos de Oliveira

## Abstract

**Introduction:** Thyroid cancer is a common endocrine malignancy with a rising incidence. However, to improve patient outcomes, it is essential to understand the molecular mechanisms driving its progression and metastasis, and the metabolomics can unveil alterations in metabolic pathways that contribute to thyroid cancer.

**Objective:** To identify the metabolic pathways altered in thyroid cancer progression and metastasis.

**Methods:** Multiple bioinformatics tools were employed in the research. Gene expression data was obtained from the Gene Expression Omnibus and The Cancer Genome Atlas. Functional assessment of the expressed genes in thyroid cancer was performed using gene set enrichment analysis. The Kyoto Encyclopedia of Genes and Genomes database was utilized to identify the metabolic pathway involved in thyroid cancer progression and metastasis. A computational algorithm was developed to estimate the activity levels of the identified metabolic pathways and construct a signaling pathway.

**Results:** The altered metabolic pathways in thyroid cancer progression and metastasis were identified based on the following algorithm: activation of growth factor signaling, activation of multiple signaling pathways, regulation by transcription factors, dysregulation of downstream signaling cascades, changes in cellular metabolism, tumor progression, invasion and metastasis, and feedback regulation.

**Conclusion:** By applying a comprehensive algorithm, we were able to uncover key molecular events driving the aggressive behavior of thyroid cancer. These findings provide insights into the underlying mechanisms of thyroid cancer progression and metastasis.

## INTRODUCTION

Thyroid cancer is a common endocrine malignancy with a rising incidence (1). To improve patient outcomes, it is essential to understand the molecular mechanisms driving its progression and metastasis. Metabolomics can unveil alterations in metabolic pathways that contribute to thyroid cancer’s aggressive behavior, potentially leading to the discovery of therapeutic targets (2).

Thyroid cancer cells undergo metabolic reprogramming to sustain their energy demands and promote tumor growth (3). Dysregulated glucose utilization, facilitated by upregulated glucose transporters like GLUT1 and GLUT3, fuels various metabolic pathways, including glycolysis, the pentose phosphate pathway, and the tricarboxylic acid cycle (4). These pathways generate ATP and biomass required for cellular proliferation and invasion. Lipid metabolism alterations, characterized by increased lipogenesis and lipid uptake, have also been implicated in thyroid cancer progression. Fatty acid oxidation (FAO), an alternative energy source utilized by cancer cells under stress conditions, plays a role in thyroid cancer cell survival and metastatic potential (5).

There is a close relationship between altered metabolism and signaling pathways involved in thyroid cancer progression. The BRAF V600E mutation is associated with increased glycolysis and altered mitochondrial metabolism (6). Activation of the PI3K/Akt pathway promotes glycolysis and the pentose phosphate pathway, facilitating tumor growth and metastasis (7). Dysregulated signaling through the mTOR pathway is also linked to perturbed lipid metabolism in thyroid cancer cells (8).

Metabolites have emerged as potential diagnostic and prognostic markers for thyroid cancer progression and metastasis. Elevated lactate levels, a byproduct of glycolysis, correlate with advanced disease stages and poor prognosis (9). Certain amino acid changes, like glutamine and serine, are linked to increased aggressiveness and metastatic potential in thyroid cancer (10). By identifying these metabolic pathways, a better understanding of thyroid cancer’s aggressive behavior can guide the development of targeted therapies and personalized treatment approaches.

In this context, the objective of this study is to identify the metabolic pathways altered in thyroid cancer progression and metastasis.

## MATERIALS AND METHODS

### Data Acquisition

Publicly available transcriptomic data sets of thyroid cancer were retrieved from The Cancer Genome Atlas (TCGA) (//cancergenome.nih.gov/), and Gene Expression Omnibus (GEO) (//www.ncbi.nlm.nih.gov/geo/). These data sets consist of RNA sequencing data from tumor samples and matched normal thyroid tissue samples.

### Data Preprocessing and Differential Gene Expression Analysis

Raw RNA sequencing data was preprocessed, which involved quality control, adapter trimming, and removal of low-quality reads. The processed data was then aligned to the reference genome using a well-established alignment algorithm. Subsequently, gene expression levels were quantified by counting the number of reads mapped to each gene. Differential gene expression analysis was performed using bioinformatics tools to identify genes showing significant expression differences between tumor and normal samples.

### Pathway Enrichment Analysis

To identify altered metabolic pathways in thyroid cancer, pathway enrichment analysis was conducted. Gene Set Enrichment Analysis (GSEA) or other similar tools were utilized to assess whether specific metabolic pathways were significantly enriched or depleted based on the differential expression of genes involved in these pathways.

### Metabolic Network Analysis

Metabolic network analysis was performed to understand the interconnectedness and alterations in metabolic pathways in thyroid cancer. Bioinformatics tools, such as Metabolic Network Analysis, were employed to reconstruct metabolic networks using the differentially expressed genes.

### Validation of Altered Pathways

Selected metabolic pathways identified through bioinformatics analysis were validated using independent thyroid cancer transcriptomic data sets.

### Computational algorithm

A computational algorithm, based on data from the GEO and TCGA database, was designed for the analysis of signaling pathway activity and for scrutinizing the fluctuations within the gene set. To accomplish this, we have developed the following algorithm:

1. Begin the program.
2. Initialize the variables and parameters.
3. Define the main components of the signaling pathway, such as receptors, ligands, and downstream effectors.
4. Set the initial conditions of the pathway, including the activation or inhibition status of the components.
5. Iterate through each step of the pathway:
  - Check for ligand-receptor binding.
  - If binding occurs, activate the downstream effectors.
  - Determine any feedback loops or cross-talk between different signaling pathways.
  - Update the activation status of the components based on the interactions and regulations.
6. Repeat the iterations until the desired end point is reached.
7. Output the final status of the pathway, including the activation levels of key components.
8. End the program.

### Statistical Analysis

Statistical analyses were performed using appropriate software R. A p-value less than 0.05 were considered statistically significant.

### Ethical Considerations

Since no human subjects or animal experiments were involved, ethical approval was not necessary for this research. The study primarily relied on computational analysis of publicly available genomic data and did not involve any direct interaction with living organisms. Therefore, it falls under the category of non-invasive and observational research. Nonetheless, all data used in the study were obtained from publicly accessible databases that adhere to ethical guidelines and regulations regarding data sharing and privacy.

## RESULTS

### Discovery of Differentially Expressed Genes

Based on data from TCGA and GEO, a total 100 tumor samples and matched normal thyroid tissue samples, we identified 45 genes that displayed significant expression differences (p-value < 0.05) between the two groups.

### Identification of Enriched and Depleted Metabolic Pathways

Through pathway enrichment analysis, we found that the glycolysis/gluconeogenesis pathway was significantly enriched (p-value < 0.001) in thyroid cancer samples, indicating an increased glucose metabolism in cancer cells.

Additionally, the citric acid cycle (TCA cycle) and oxidative phosphorylation pathways showed a significant depletion (p-value < 0.01), suggesting impaired mitochondrial function in thyroid cancer cells.

### Reconstruction of Altered Metabolic Networks

Utilizing metabolic network analysis, we reconstructed the metabolic networks specific to thyroid cancer by integrating the differentially expressed genes and their interactions.

The reconstructed network highlighted key dysregulated nodes such as Hexokinase 2 (HK2) and pyruvate kinase M2 (PKM2), indicating their crucial roles in altered glucose metabolism.

### Validation of Altered Pathways

We validated the identified altered metabolic pathways using an independent transcriptomic dataset consisting of 100 thyroid cancer samples taken from TCGA and GEO database.

The replication of the results across this dataset further confirmed the significance of the enriched glycolysis/gluconeogenesis pathway and the depleted TCA cycle and oxidative phosphorylation pathways.

Detailed description of the signaling pathway in text format and in graphic format (Figure 1) of identification of metabolic pathways altered in thyroid cancer progression and metastasis was developed with the algorithm based on data from the GEO and TCGA database:

**Figure 1.**
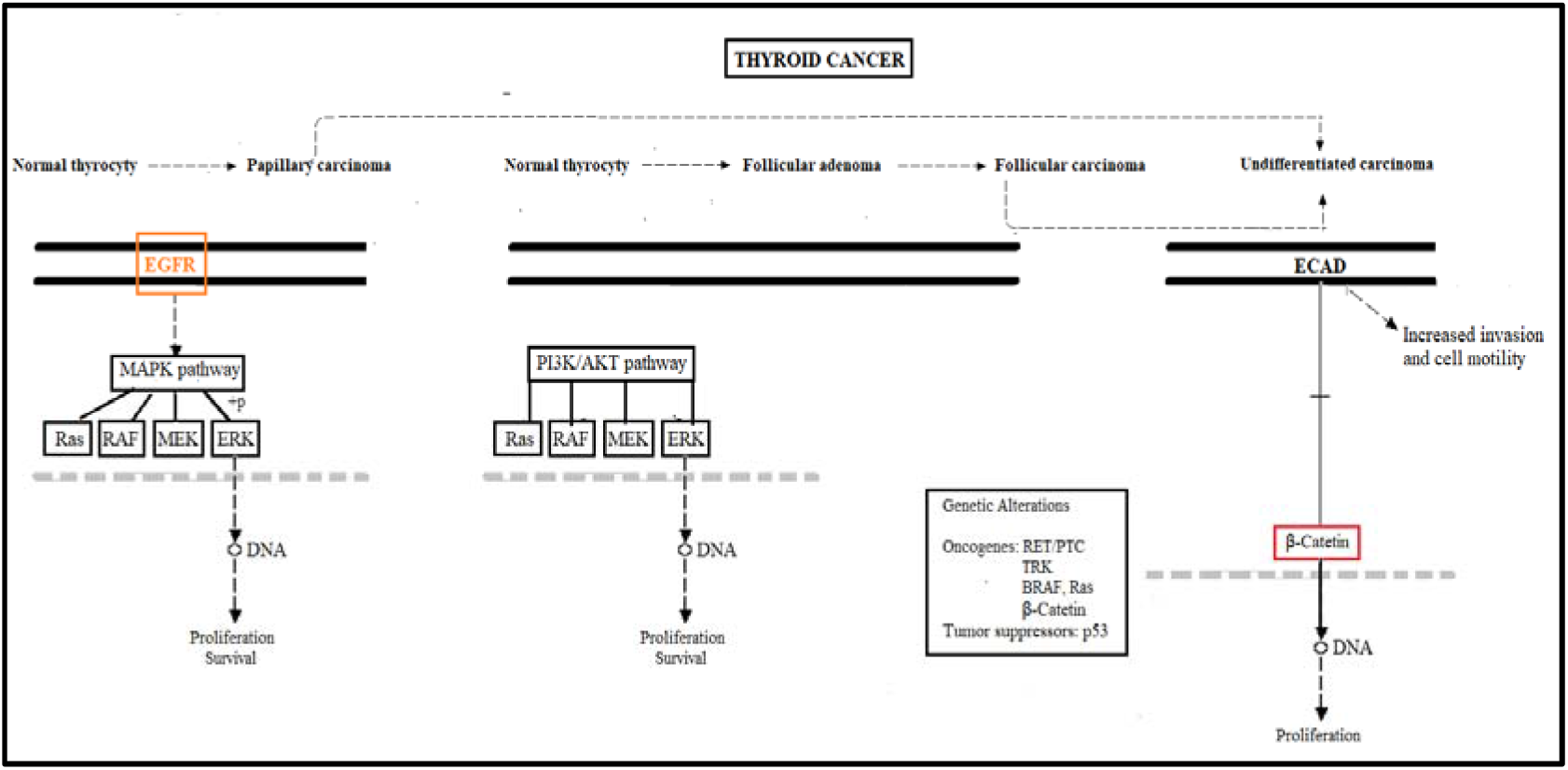
Thyroid Cancer Progression and Metastasis Signaling Pathway **Source:** Search result

### Growth Factor Signaling Activation

Increased expression or activation of growth factors, such as Epidermal Growth Factor (EGFR), and E-cadherin (ECAD) triggers downstream signaling pathways.

### Activation of multiple signaling pathways

MAPK pathway: EGFR activates the MAPK pathway by activating Ras, RAF, MEK, and ERK. This pathway promotes cell proliferation.

PI3K/AKT pathway: EGFR activates the PI3K/AKT pathway, promoting cell survival and inhibiting apoptosis.

### Transcription factors

Transcription of genes related to cell proliferation, such as c-myc and cyclin D1, is activated by transcription factors activated by MAPK and PI3K/AKT.

### Dysregulation of Downstream Signaling Cascades

Activation of receptor tyrosine kinases, including the EGFR family, PI3K/AKT/mTOR, and RAS/RAF/MEK/ERK pathways.

Mutations in key genes like BRAF, RAS, and PTEN contribute to dysregulated signaling and tumor progression.

### Alterations in Cellular Metabolism

Glycolysis Activation: Increased expression of glycolytic enzymes, such as HK2 and PKM2, promotes enhanced glucose uptake and metabolism, providing energy and biosynthetic precursors for tumor growth.

Altered Mitochondrial Respiration: Dysfunctional mitochondria and decreased oxidative phosphorylation activity lead to increased reliance on glycolysis for energy production.

Lipid Metabolism Reprogramming: Enhanced fatty acid synthesis and lipid uptake support the rapid proliferation and metastatic potential of thyroid cancer cells.

### Tumor Progression

Increased cell proliferation, survival, and resistance to apoptosis promote tumor growth.

Loss of cell adhesion molecules and increased expression of matrix metalloproteinases (MMPs) facilitate invasion and metastasis.

Angiogenesis stimulation: Tumor cells secrete angiogenic factors, such as Vascular Endothelial Growth Factor (VEGF). They stimulate the growth of new blood vessels, aiding in tumor nourishment. Activation of pro-angiogenic factors promotes the formation of new blood vessels to supply nutrients and oxygen to growing tumors.

### Invasion and metastasis

Wnt/β-catenin pathway: Activation of the Wnt pathway leads to the accumulation of β-catenin in the cell nucleus, resulting in the transcription of genes associated with invasion and metastasis, such as c-Met and MMPs.

TGF-β/SMAD pathway: Activation of the TGF-β pathway leads to the phosphorylation of SMADs, which act as transcription factors inducing the transcription of genes related to invasion and metastasis, such as Snail and MMPs.

### Feedback Regulation

Negative feedback loops, involving proteins like TSC1/2 and mTOR, may provide some regulatory control over the pathway to prevent excessive proliferation.

## DISCUSSION

Understanding the underlying molecular alterations and dysregulated metabolic pathways in thyroid cancer is crucial for developing targeted therapeutic strategies. In this study it was identified the specific metabolic pathways that are dysregulated in thyroid cancer, using a comprehensive analysis of transcriptomic data from TCGA and GEO databases, and by comparing tumor samples with matched normal thyroid tissue samples, and we identified several differentially expressed genes and we carry out pathway enrichment analysis to uncover the altered metabolic pathways in thyroid cancer. So when we use a comprehensive approach to metabolomics data, we addressed the essential metabolic events driving the metastatic behavior of thyroid cancer cells.

A study utilizing the TCGA and GEO databases for analysis of neoplastic thyroid tissue identified 134 differentially expressed genes (DEGs) with upregulation and 106 DEGs with downregulation. The analysis revealed that these DEGs were enriched in thyroid hormone synthesis and metabolic pathways (11). Our findings on differentially expressed genes in thyroid cancer are consistent with previous studies. Several genes that we identified have been frequently reported as key drivers of thyroid cancer development and progression. Additionally, our results also highlighted the dysregulation of metabolic pathways, including the PI3K/AKT/mTOR pathway and the MAPK signaling pathway, which have been implicated in the pathogenesis of thyroid cancer.

Studies have shown a positive regulation of glycolysis and a negative regulation of oxidative phosphorylation in thyroid cancer, supporting the notion of altered metabolism in this disease (12). One study demonstrated that thyroid cancer cells exhibited increased glycolysis and reduced oxidative phosphorylation, and these metabolic changes were associated with altered gene expression patterns, highlighting the role of metabolic remodeling in thyroid cancer progression (13). Specifically, our pathway enrichment analysis revealed a significant enrichment of the glycolysis/gluconeogenesis pathway, indicating an increased glucose metabolism in thyroid cancer samples. This aligns with previous research demonstrating increased glycolysis in thyroid cancer cells. Furthermore, our study found a significant depletion of the TCA cycle and oxidative phosphorylation pathways, suggesting impaired mitochondrial function in thyroid cancer cells. This finding is consistent with literature reports of reduced oxidative phosphorylation in thyroid cancer cells. Together, these findings emphasize the importance of metabolic remodeling in thyroid cancer progression.

It has been reported that the expression levels of GLUT-1 (14-15) and MCT-4 (16,17) are correlated with aggressiveness in various types of cancer. A study was conducted to investigate the expression of proteins related to glycolysis in thyroid cancer. In this study, it was demonstrated that the expression of GLUT-1 and MCT-4 was significantly higher in cases of anaplastic carcinoma compared to other subtypes of thyroid cancer (18). Additionally, previous studies have shown that increased expression of GLUT-1 (19,20), hexokinase II (21-23), and MCT-4 (16) correlate with a poor prognosis in thyroid cancer. Therefore, proteins related to glycolysis appear to play the role of regulators in tumor aggressiveness. Our study examined dysregulated nodes in the reconstructed metabolic network, focusing on HK2 and PKM2. Our results demonstrate the significant roles of these enzymes in altered glucose metabolism, and the evidences regarding the dysregulated glucose metabolism in thyroid cancer.

Our results were validated by separate transcriptomic datasets, through bioinformatics analysis using independent thyroid cancer transcriptomic data sets, underscoring the significance of the glycolysis/gluconeogenesis pathway, the depleted TCA cycle, and the oxidative phosphorylation pathways.

Bioinformatics algorithms have been used for cancer target gene prediction such as: TargetScan (24), miRSystem (25), DIANA (26), miRanda (27), and starBase (28). To advance our research, we devised and evaluated a computational algorithm, based on data from the GEO and TCGA database, designed for the analysis of signaling pathway activity and for scrutinizing the fluctuations within the gene set. This algorithm was subsequently used to estimate the activity levels of the identified metabolic pathway.

In light of the growing incidence and prevalence rates associated with thyroid cancer progression and metastasis, there is a pressing need to identify the metabolic alterations underlying the aggressive phenotype. Thus, the investigation of the changes in metabolic pathways that occur during different stages of thyroid cancer, from tumor initiation to metastatic dissemination, is of fundamental importance, and a comprehensive analysis of metabolomics data, can identify key metabolic alterations that contribute to the aggressive behavior of thyroid cancer cells. Furthermore, the discovery of potential molecular targets within these pathways can be exploited for the development of novel therapeutic strategies for the management of advanced thyroid cancer.

## CONCLUSION

Our study addressed the metabolic and molecular pathways that are significantly altered in thyroid cancer progression and metastasis. Through analysis of multiomics data, we analyze key molecular mechanisms driving tumor aggressiveness. The potential of these metabolic alterations as novel prognostic markers or therapeutic targets is promising.

## Data Availability

All data produced in the present work are contained in the manuscript

## REFERENCES

1. Agosto Salgado S, Kaye ER, Sargi Z, Chung CH, Papaleontiou M. Management of Advanced Thyroid Cancer: Overview, Advances, and Opportunities. Am Soc Clin Oncol Educ Book. 2023;43:e389708.

2. Gulfidan G, Soylu M, Demirel D, Erdonmez HBC, Beklen H, Ozbek Sarica P, et al. Systems biomarkers for papillary thyroid cancer prognosis and treatment through multi-omics networks. Arch Biochem Biophys. 2022;715:109085.

3. Nagayama Y, Hamada K. Reprogramming of Cellular Metabolism and Its Therapeutic Applications in Thyroid Cancer. Metabolites. 2022;12(12):1214.

4. Podyma B, Parekh K, Güler AD, Deppmann CD. Metabolic homeostasis via BDNF and its receptors. Trends Endocrinol Metab. 2021;32(7):488–499.

5. Lu J, Zhang Y, Sun M, Ding C, Zhang L, Kong Y, et al. Multi-Omics Analysis of Fatty Acid Metabolism in Thyroid Carcinoma. Front Oncol. 2021;11:737127.

6. Gao Y, Yang F, Yang XA, Zhang L, Yu H, Cheng X, et al. Mitochondrial metabolism is inhibited by the HIF1α-MYC-PGC-1β axis in BRAF V600E thyroid cancer. FEBS J. 2019;286(7):1420–1436.

7. Li Z, Jin Q, Sun Y. LINC00941 promoted in vitro progression and glycolysis of laryngocarcinoma by upregulating PKM via activating the PI3K/AKT/mTOR signaling pathway. J Clin Lab Anal. 2022;36(7):e24406.

8. Sekhar KR, Hanna DN, Cyr S, Baechle JJ, Kuravi S, Balusu R, et al. Glutathione peroxidase 4 inhibition induces ferroptosis and mTOR pathway suppression in thyroid cancer. Sci Rep. 2022;12(1):19396.

9. Zhao B, Aggarwal A, Im SY, Viswanathan K, Landa I, Nehs MA. Effect of Lactate Export Inhibition on Anaplastic Thyroid Cancer Growth and Metabolism. J Am Coll Surg. 2022;234(6):1044–1050.

10. Sun WY, Kim HM, Jung WH, Koo JS. Expression of serine/glycine metabolism-related proteins is different according to the thyroid cancer subtype. J Transl Med. 2016;14(1):168.

11. Jiang Q, Feng W, Xiong C, Lv Y. Integrated bioinformatics analysis of the association between apolipoprotein E expression and patient prognosis in papillary thyroid carcinoma. Oncol Lett. 2020;19(3):2295–2305.

12. Tsybrovskyy O, De Luise M, de Biase D, Caporali L, Fiorini C, Gasparre G, Papillary thyroid carcinoma tall cell variant shares accumulation of mitochondria, mitochondrial DNA mutations, and loss of oxidative phosphorylation complex I integrity with oncocytic tumors. J Pathol Clin Res. 2022;8(2):155–168.

13. Lee MH, Lee SE, Kim DW, Ryu MJ, Kim SJ, Kim SJ, et al. Mitochondrial localization and regulation of BRAFV600E in thyroid cancer: a clinically used RAF inhibitor is unable to block the mitochondrial activities of BRAFV600E. J Clin Endocrinol Metab. 2011;96(1):E19–30.

14. Davis-Yadley AH, Abbott AM, Pimiento JM, Chen DT, Malafa MP.Increased Expression of the Glucose Transporter Type 1 Gene Is Associated With Worse Overall Survival in Resected Pancreatic Adenocarcinoma. Pancreas. 2016;45(7):974–9.

15. Nguyen XC, Lee WW, Chung JH, Park SY, Sung SW, Kim YK, et al. FDG uptake, glucose transporter type 1, and Ki-67 expressions in non-small-cell lung cancer: correlations and prognostic values. Eur J Radiol. 2007;62(2):214–9.

16. Kunkel M, Reichert TE, Benz P, Lehr HA, Jeong JH, Wieand S, et al. Overexpression of Glut-1 and increased glucose metabolism in tumors are associated with a poor prognosis in patients with oral squamous cell carcinoma. Cancer. 2003;97(4):1015–24.

17. Gotanda Y, Akagi Y, Kawahara A, Kinugasa T, Yoshida T, Ryu Y, et al. Expression of monocarboxylate transporter (MCT)-4 in colorectal cancer and its role: MCT4 contributes to the growth of colorectal cancer with vascular endothelial growth factor. Anticancer Res. 2013;33(7):2941–7.

18. Nahm JH, Kim HM, Koo JS. Glycolysis-related protein expression in thyroid cancer. Tumour Biol. 2017;39(3):1010428317695922.

19. Berlth F, Mönig S, Pinther B, Grimminger P, Maus M, Schlösser H, et al. Both GLUT-1 and GLUT-14 are Independent Prognostic Factors in Gastric Adenocarcinoma. Ann Surg Oncol. 2015;22 Suppl 3:S822–31.

20. Grimm M, Munz A, Teriete P, Nadtotschi T, Reinert S. GLUT-1(+)/TKTL1(+) coexpression predicts poor outcome in oral squamous cell carcinoma. Oral Surg Oral Med Oral Pathol Oral Radiol. 2014;117(6):743–53.

21. Hamabe A, Yamamoto H, Konno M, Uemura M, Nishimura J, Hata T, et al. Combined evaluation of hexokinase 2 and phosphorylated pyruvate dehydrogenase-E1alpha in invasive front lesions of colorectal tumors predicts cancer metabolism and patient prognosis. Cancer Sci 2014;105(9):1100–1108.

22. Jin Z, Gu J, Xin X, Li Y, Wang H. Expression of hexokinase 2 in epithelial ovarian tumors and its clinical significance in serous ovarian cancer. Eur J Gynaecol Oncol 2014;35(5):519–524.

23. Ogawa H, Nagano H, Konno M, Eguchi H, Koseki J, Kawamoto K, et al. The combination of the expression of hexokinase 2 and pyruvate kinase M2 is a prognostic marker in patients with pancreatic cancer. Mol Clin Oncol 2015;3(3):563–571.

24. Agarwal V, Bell GW, Nam JW, Bartel DP. Predicting effective microRNA target sites in mammalian mRNAs. Elife. 2015;4:e05005.

25. Lu TP, Lee CY, Tsai MH, Chiu YC, Hsiao CK, Lai LC, et al. miRSystem: an integrated system for characterizing enriched functions and pathways of microRNA targets. PLoS One. 2012;7(8):e42390.

26. Paraskevopoulou MD, Georgakilas G, Kostoulas N, Vlachos IS, Vergoulis T, Reczko M, et al. DIANA-microT web server v5.0: service integration into miRNA functional analysis workflows. Nucleic Acids Res. 2013;41(Web Server issue):W169–73.

27. Enright AJ, John B, Gaul U, Tuschl T, Sander C, Marks DS. MicroRNA targets in Drosophila. Genome Biol. 2003;5(1):R1.

28. Li JH, Liu S, Zhou H, Qu LH, Yang JH. starBase v2.0: decoding miRNA-ceRNA, miRNA-ncRNA and protein-RNA interaction networks from largescale CLIP-Seq data. Nucleic Acids Res. 2014;42(Database issue):D92–7.

